# Putamen dopamine synthesis, vesicular storage, and metabolism in Parkinson disease

**DOI:** 10.64898/2026.03.12.26348277

**Authors:** David S. Goldstein

**Affiliations:** Clinical Neurosciences Program, Division of Intramural Research, National Institute of Neurological Disorders and Stroke, National Institutes of Health, Bethesda, MD

## Abstract

**Background:** Severe putamen dopamine depletion in Parkinson disease (PD) has been attributed to nigrostriatal denervation; however, there are also functional abnormalities in extant terminals (the “sick-but-not-dead” phenomenon). Rates of intra-neuronal processes of synthesis, storage, and metabolism of dopamine complexly influence releasable dopamine stores but have not yet been systematically estimated.

**Methods:** Post-mortem empirical data were available about putamen tissue contents of 7 reactants, including the autotoxic dopamine metabolite 3,4-dihydroxyphenylacetaldehyde (DOPAL). We constructed kinetic models depicting reactions related to putamen dopamine content, the simplest model consisting of 7 reactions and the most complete model 18 reactions among 10 intra-neuronal reactants. We used the post-mortem data, in vivo results of ^18^F-DOPA positron emission tomography (PET), and the models to estimate rates of the intra-neuronal processes and rank their contributions to control-PD differences.

**Results:** There was about a 98% decrease in putamen tissue dopamine in PD. The concentration ratio of DOPAL/DA was about 9 times control. Applying the simplest kinetic model, vesicular sequestration was estimated to be decreased by 98.5% (0.073 vs. 4.91 nmol/min). About 3-fold greater in vivo “washout” of putamen ^18^F-DOPA-derived radioactivity compared to controls also indicated attenuated vesicular storage in PD. According to the complete model, control-PD differences in intra-neuronal reaction rates were, in descending order, vesicular uptake ≈ vesicular leakage > exocytotic release ≈ neuronal reuptake > L-aromatic-amino-acid decarboxylase activity ≈ tyrosine hydroxylase activity > other reactions.

**Discussion:** Empirical post-mortem and in vivo data and application of kinetic models provide convergent quantitative evidence for a substantial vesicular storage defect in residual dopaminergic terminals in PD, a potential target for disease-modifying treatment or prevention strategies.

**Trial Registration:** None

**BRIEF SUMMARY:** We estimated rates of reactions involved with synthesis, storage, release, reuptake, and metabolism of dopamine in the putamen in Parkinson disease and found that the main intra-neuronal functional abnormality separating Parkinson disease from controls was attenuated vesicular sequestration, implicating decreased vesicular uptake via the vesicular monoamine transporter and increased vesicular leakiness as key determinants of putamen dopamine deficiency in PD.

## INTRODUCTION

The movement disorder that characterizes Parkinson disease (PD) is associated with and largely results from profound striatal dopamine deficiency.^1^ Post-mortem studies have shown that the putamen is the striatal component with the most severe dopamine depletion—about 98%. ^2, 3^ This drastic central neurochemical lesion is the proximate cause of the movement disorder, since bypassing the defect with levodopa/carbidopa treatment often dramatically improves motor symptoms of PD.^4^

Although PD has been attributed to loss of dopaminergic neurons in the substantia nigra pars compacta,^5^ evidence from imaging, neurochemical, and post-mortem studies indicates that the dopamine deficiency reflects not only loss of nigrostriatal innervation but also functional impairments in surviving nerve terminals, such as reduced dopamine synthesis, attenuated vesicular storage and release, and increased cytosolic dopamine oxidation.^6, 7^ These abnormalities may be in operation before loss of nigral melanized neurons.^8^

Consistent with this view, computational modeling has revealed that after taking denervation into account multiple functional abnormalities are apparent in the synthesis, storage, reuptake, and metabolism of catecholamines in cardiac sympathetic noradrenergic nerves in PD^9^—especially a shift from vesicular sequestration to enzymatic oxidative deamination of cytoplasmic catecholamines.^10^ Analogous intra-neuronal abnormalities might also be present in the putamen; however, this possibility has not yet been systematically assessed.

To fill this knowledge gap we constructed a series of increasingly complex kinetic models that depict the known reactions and reactants involved with putamen vesicular dopamine stores. The simplest model (Figure 1A) was expanded in a step-wise manner primarily by adding reactions with relatively small rates relative to the dominant fluxes. The final model consisted of 18 reactions among 9 reactants (Figure 2).

**Figure 1:**
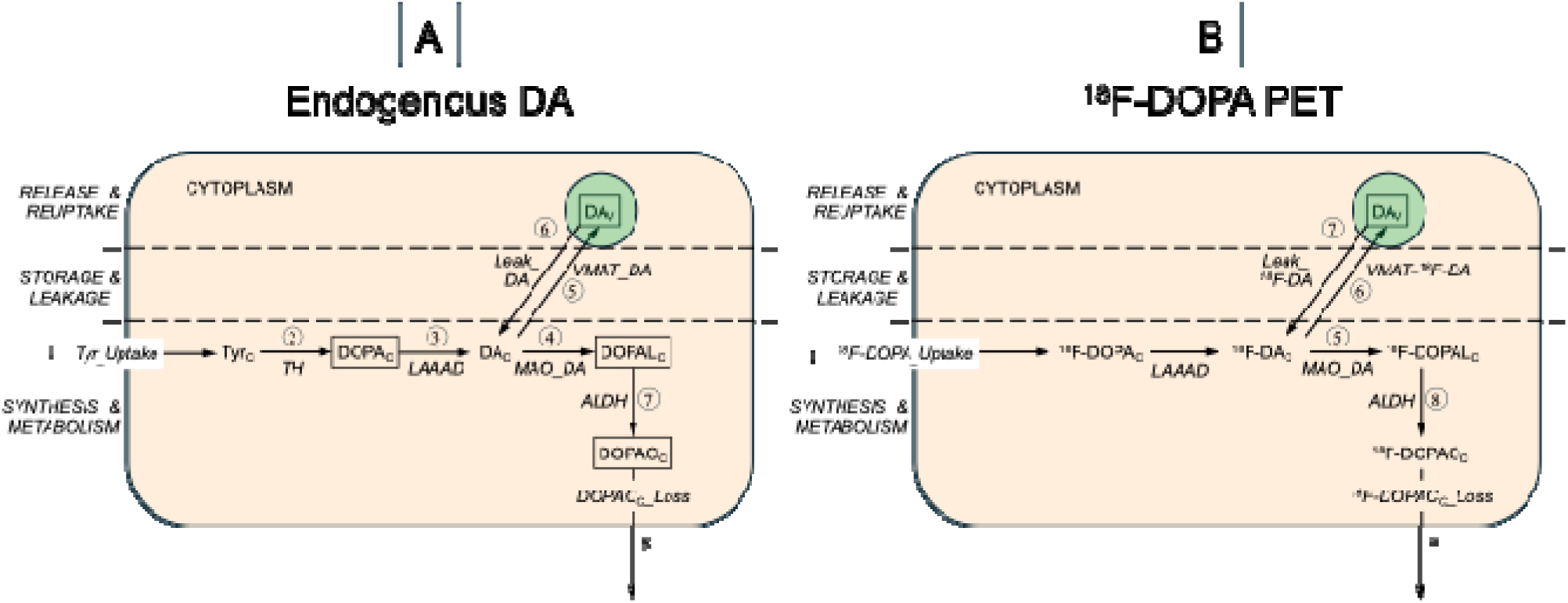
Initial model of (A) putamen endogenous DA synthesis, storage, and turnover and (B) putamen ^18^F-DOPA-derived radioactivity. The model in (A) does not include release and reuptake of vesicular dopamine (DAv), on the grounds that under resting conditions the main determinant of tissue DA turnover is metabolism of cytoplasmic DA (DAc) by monoamine oxidase (MAO_DA) to form 3,4-dihydroxyphenylacetaldehyde in the cytoplasm (DOPALc), followed by enzymatic oxidation of DOPAL via aldehyde dehydrogenase (ALDH) to form 3,4-dihydroxyphenylacetic acid (DOPACc) and then rapid extrusion of DOPACc from the terminal (DOPACc_Loss). There are 8 reactions (circled numbers); there are 4 intra-neuronal reactants for which empirical post-mortem data are available in PD and control groups (abbreviations in rectangles). The intra-neuronal reactions are: (1) Tyr_Uptake = rate of uptake of tyrosine; (2) TH = rate of enzymatic hydroxylation of tyrosine by tyrosine hydroxylase; (3) LAAAD = rate of enzymatic decarboxylation of cytoplasmic DOPA (DOPAc); (4) MAO_DA=MAO-mediated DA enzymatic oxidation (conversion of DAc to DOPALc); (5) VMAT_DA = rate of vesicular uptake of cytoplasmic DA; (6) Leak_DA = rate of passive leakage of DA from the vesicles into the cytoplasm; (7) ALDH = rate of metabolism of DOPALc to DOPACc catalyzed by aldehyde dehydrogenase; (8) DOPACc_Loss = rate of loss of DOPACc from the model. The model in (B) shows corresponding reactions after administration of ^18^F-DOPA to visualize putamen dopaminergic innervation by positron emission tomography (PET).

**Figure 2:**
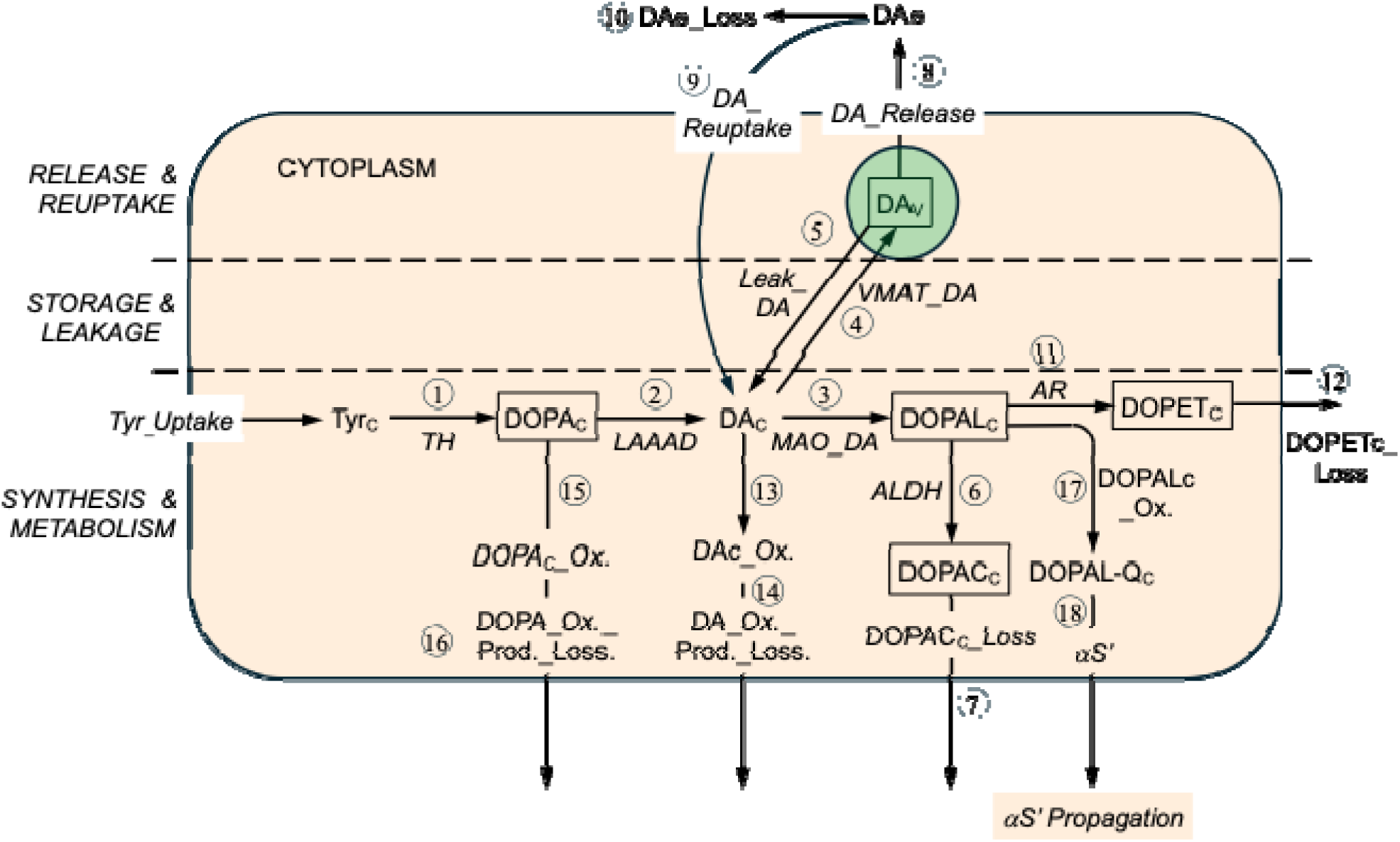
Final model. There are 18 reactions (circled numbers) and intra-neuronal reactants for which empirical post-mortem data are available in PD and control groups (abbreviations in rectangles). The intra-neuronal reactions are: ALDH=aldehyde dehydrogenase; AR=aldehyde reductase; DA_Release=exocytotic release of vesicular dopamine; DA_Reuptake=reuptake of dopamine from the extracellular fluid; DAc_Ox.=spontaneous oxidation of cytoplasmic dopamine; DOPAc_Ox._Prod._Loss= loss of spontaneous oxidation products of cytoplasmic DOPA from the model; DOPAc_Ox.=spontaneous oxidation of cytoplasmic DOPA; DOPACc_Loss=loss of cytoplasmic 3,4-dihydroxyphenylacetic acid from the model; DOPALc_Ox.=spontaneous oxidation of cytoplasmic 3,4-dihydroxyphenylacetaldehyde; DOPETc_Loss=loss of cytoplasmic 3,4-dihydroxyphenylethanol from the model; LAAAD=L-aromatic-amino-acid decarboxylase; Leak_DA=leakage of vesicular dopamine into the cytoplasm; MAO_DA=monoamine oxidase acting on cytoplasmic dopamine; TH=tyrosine hydroxylase; VMAT_DA=vesicular monoamine transporter acting on cytoplasmic dopamine; αS’ Formation=formation of modified alpha-synuclein

Post-mortem empirical data were available about putamen tissue contents of 7 of the reactants, including the autotoxic dopamine metabolite 3,4-dihydroxyphenylacetaldehyde (DOPAL).^3, 11^ For in vivo verification we constructed a similar model of putamen ^18^F-DOPA-derived radioactivity quantified by positron emission tomography (PET) (Figure 1B).

We applied the final model to the post-mortem neurochemical and in vivo PET data and compared the results to previously published findings from similar assessments of reactions in cardiac sympathetic nerves.^12^ Our hypothesis was that the same intra-neuronal functional abnormalities identified previously in the heart would be found in the putamen in PD.

## METHODS

### Study approval

Data retrieval and analyses for this project were conducted under a secondary use protocol approved by the Institutional Review Board of the National Institutes of Health (NIH), Clinical Protocol 000490, “Mechanisms of Autonomic and Catecholamine-Related Disorders.”

Post-mortem data were culled from an ongoing database of samples assayed at the NIH from autopsies done at the NIH Clinical Center or at outside centers that had provided tissue samples. In all cases, written informed consent had been obtained from the next of kin in advance of the tissue harvesting. Most of the data have been published previously.^3, 6, 13, 14^ The autopsies were performed from 1996 to 2024.

Published concentrations in units of ng/mg protein were converted to concentrations per mg wet weight based on protein being 12% of the wet weight. Putamen concentrations per unit wet weight were brought to reactant amounts assuming tissue putamina volume totaling 10 mL in the Control and PD groups.

### Initial model corresponding to empirical ^18^F-DOPA data

Of the intra-neuronal reactants in the simplest model, empirical post-mortem data were available for 4—cytosolic DOPA (DOPAc), DOPAL (DOPALc), 3,4-dihydroxyphenylacetic acid (DOPACc), and vesicular dopamine (DAv), assuming all of intra-neuronal DOPA, DOPAL, and DOPAC is in the cytosol and dopamine in vesicles. Relevant to the estimated rate of “net” vesicular sequestration based on analysis of ^18^F-DOPA PET data, Figure 1B depicts the model adapted for the uptake, storage, and metabolism of ^18^F-DOPA in the putamen.

The simplest model consisted of 8 reactions (circled numbers in Figure 1) and 4 intra-neuronal reactants (abbreviations in rectangles in Figure 1). The intra-neuronal reactions in this model were: (1) Tyr_Uptake = rate of uptake of tyrosine; (2) TH = rate of enzymatic hydroxylation of tyrosine by tyrosine hydroxylase; (3) LAAAD = rate of enzymatic decarboxylation of cytoplasmic DOPA (DOPAc); (4) MAO_DA=MAO-mediated DA enzymatic oxidation (conversion of DAc to DOPALc); (5) VMAT_DA = rate of vesicular uptake of cytoplasmic dopamine; (6) Leak_DA = rate of passive leakage of dopamine from the vesicles into the cytoplasm; (7) ALDH = rate of metabolism of DOPALc to DOPACc catalyzed by aldehyde dehydrogenase; (8) DOPACc_Loss = rate of loss of DOPACc from the model. The model in Figure 1B shows the corresponding reactions after administration of ^18^F-DOPA to visualize putamen dopaminergic innervation by PET.

We refined the model in a step-by-step fashion, as follows.

### Refinement 1: Release, reuptake, and loss of dopamine (alternative to loss by intra-neuronal metabolism)

Under resting conditions, most of the turnover of endogenous catecholamine occurs by metabolism in the cytoplasm, which is about 3-7 times that of exocytotic release with escape from neuronal reuptake^15, 16^ The rate of exocytotic release of dopamine (and consequently the rates of neuronal reuptake and of loss of extracellular fluid dopamine from the model) therefore normally is small with respect to the rate of MAO acting on DAc.

Nevertheless, in PD putamen there is almost a complete absence of DAT sites,^17^ and across individuals putamen DAT protein is strongly positively correlated with tissue dopamine content.^18, 19^ Because of these considerations, exocytotic release of dopamine with escape of reuptake should not be presumed to be negligible. The model therefore was refined to include DA_Release, DA_Reuptake, and loss of extracellular fluid dopamine from the model (DAe_Loss). We presumed that DA_Reuptake was 90% of DA_Release in controls^20^ and 10% of DA_Release in PD.^17–19^ By inspection of Figure 2, we adopted the following equilibrium equation.

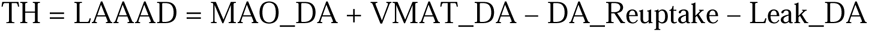

Based on available post-mortem data^21–25^ and our previous modeling of reactions in cardiac sympathetic nerves ^9^, under resting conditions only a very small percent of DAv is released per minute (0.1%-0.2%).

Using the steady-state identities together with the spreadsheet values, we computed the fluxes in each group—without needing DAc—by chaining these relations:

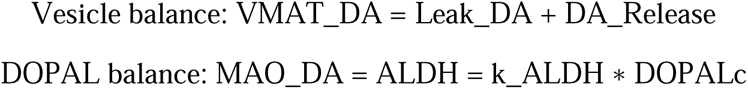

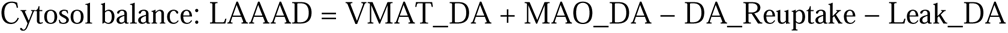

Cytosol balance: LAAAD = VMAT_DA + MAO_DA − DA_Reuptake − Leak_DA with group constraints specified for DA_Reuptake = 0.9 _*_ DA_Release in Control and 0.1 _*_ DA_Release in PD and with first-order fluxes when reactant amount were known.

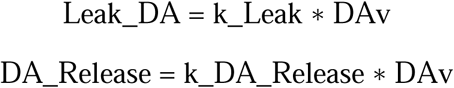

### Refinement 2: DOPET (alternative to DOPAC as a metabolite of DOPAL)

In putamen tissue from control subjects, the median concentration of 3,4-dihydroxyphenylethanol (DOPET), 0.0303 pmol/mg wet weight, is about 1% of the concentration of DOPAC (eData 1), consistent with the effective rate constant for enzymatic reduction of DOPALc by aldehyde reductase (AR) to form DOPET being very small with respect to the rate constant for enzymatic oxidation catalyzed by ALDH to form DOPAC.^26–28^

### Refinement 3: Spontaneous oxidation of cytoplasmic DA (alternative to MAO_DA)

The rate constant for enzymatic oxidation of cytoplasmic DA (DAc) by monoamine oxidase (MAO_DA) is about 7 times that for spontaneous oxidation.^29^ The effective rate constant for spontaneous oxidation of cytoplasmic DA (DA_Ox.) to form products such as 5-S-cysteinyldopamine (Cys-DA) was therefore small compared to the rate constant for enzymatic oxidative deamination catalyzed by MAO_DA to form DOPAL.

### Refinement 4: Spontaneous oxidation of cytoplasmic DOPA (alternative to LAAAD)

The rate of spontaneous oxidation of cytoplasmic DOPA (DOPAc) to form compounds such as 5-S-cysteinyldopa (Cys-DOPA) is quite small compared to the rate of enzymatic decarboxylation catalyzed by LAAAD to form dopamine. This presumption was justified, because post-mortem tissue concentrations of Cys-DOPA in control subjects were below the detection limit (data not shown), whereas tissue DOPA, dopamine, and DOPAC concentrations were always measurable. Under physiological conditions, DOPA in aqueous solution oxidizes spontaneously with a rate constant in the range of 0.001 h^−1^ (about 0.00002 min^−1^).^30^ In contrast, the rate constant for LAAAD (kLAAAD) is about 0.011 min^−1^.^9^

At steady-state the TH flux equals the sum of decarboxylation (LAAAD) and the minor rate of DOPAc oxidation (DOPAc_Ox.). Because the latter term is very small, the rate of LAAAD is about the same as the rate of TH.

### Refinement 5: Spontaneous oxidation of cytoplasmic DOPAL

A final refinement of the model related to alpha-synucleinopathy. DOPAL potently oligomerizes, forms quinoprotein adducts with, and precipitates alpha-synuclein^31, 32^ and also misfolds numerous other intracellular proteins. The mediator of these effects seems to DOPAL-quinone, which is formed from the spontaneous oxidation of DOPAL.^33–36^

When MO3.13 oligodendrocytes are incubated with DOPAL, quinones accumulate over the course of about 24 hours,^32^ whereas the rate constant for ALDH is in the range of about 0.03-0.06 min^−1^.^37^ The spontaneous oxidation rate of cytoplasmic DOPAL to form DOPAL-quinone (DOPALc_Ox.) therefore is very small compared to the rate of enzymatic oxidation of DOPALc by ALDH to form cytoplasmic DOPAC. Based on a half-time of accumulation of DOPAL-Q of about 500 min^35^ under non-oxidative conditions, a reasonable effective rate constant would be in the range of about 0.001 min^−1^.^35^

In the model, modification of alpha-synuclein depends on DOPAL-Q, since anti-oxidation with N-acetylcysteine mitigates DOPAL-induced alpha-synuclein oligomerization and quinoprotein adduct formation.^32^ The model includes DOPALc_Ox. because of the implications for formation and propagation of misfolded alpha-synuclein. The final listing of intra-neuronal reactions is in Table 1.

### Assumptions about reaction rates

For purposes of calculating reaction rates the model treated the PD state as if it were in equilibrium. It was reasonable to presume that over the course of minutes or hours approximately steady-state conditions would obtain, in contrast with progressive changes over the course of millions of minutes in a person’s lifetime (there are about ½ million minutes in a year).

All the modeled reactions were also assumed to follow first-order kinetics, meaning that substrate concentrations were below the enzymatic Michaelis-Menten constants (Km). For MAO_DA, the Km value is in the range of 20-50 µM, far greater than DAc;^38^ and for ALDH, the Km value is in the range of 0.4-1 µM, depending on ALDH1A1 or ALDH2,^39^ whereas the median empirical DOPALc concentration is about 0.0004 µmol/g. The Km for TH is in the range of 20-100 µM,^40^ depending importantly on phosphorylation,^41^ while human post-mortem putamen Tyr concentrations are in the range of 0.3 µmol/g (eTable 1).

It was possible that TH was operating at close to the saturating concentration of substrate. As noted below, however, in the model the rate of TH was estimated from an equilibrium equation, not from k _*_ Amount, and substrate saturation would not affect the equilibrium-based TH estimate.

Reaction rates were not directly fitted to experimental time-series data but were derived from steady-state constraints, empirical reactant amounts, and literature-based effective rate constants, yielding internally consistent estimates and reaction rate rankings rather than uniquely identifiable kinetic parameters.

For derivation and justifcation of formulas for reaction rates in the final model see eMethods Reaction Rate Equations.

### Statistical Methods

Because of the nature of the sources of empirical data on the reactants, fit statistics and residual plots were not done—only sensitivity ranks. Reaction-rate differences were model-derived effect sizes, not hypothesis-tested endpoints.

To assess the reasonableness of assigned kDA_Release and kALDH values obtained from the literature, one-way sensitivity analyses were done, increasing kDA_Release by up to 10-fold and decreasing kALDH by up to 90%.^42^

### Artificial Intelligence (AI) Usage

The authors use ChatGPT (versions 4, 5, and 5.1) via numerous prompts for culling literature citations and generating drafts of text and Tables in the Methods, Results, and Discussion sections. ChatGPT was used to generate the sensitivity analyses reported in eTable 2 an eTable 3.

### Data availability

Individual post-mortem data about putamen dopamine and related compounds are in eData 1-Autopsy Individual Data. Publications used for obtaining Reactant Amounts applied in the models are in eTable 1, Reactants in the Literature. The averaged post-mortem data used for Reactant amounts, the Reactions, and the estimated Reaction rates are in eResults, Reactants & Reaction Rates in Final Model.

## RESULTS

### Reactant Amounts

Amounts of putamen reactants were calculated based on values from an ongoing database under NIH Clinical Protocol 000490 (eData 1) and on published median values of putamen concentrations from culled post-mortem studies (eTable 1).

Although tissue DOPAL content was decreased in PD, tissue dopamine was decreased to a greater extent, so that the DOPAL/DA ratio was increased about 9-fold in PD compared to Control (eData 1).

### Estimated reaction rates

Application of the equations for reaction rates led to calculated values in the Control and PD groups (Table 1 and eResults). In the Control group, the highest-ranked reactions were those governing vesicular dopamine handling, including vesicular uptake (VMAT_DA), vesicular leak, exocytotic release, and reuptake. These processes dominated the flux hierarchy because of the large vesicular dopamine pool and the steady cycling of dopamine among vesicular, cytosolic, and extracellular compartments. Dopamine synthesis (TH/LAAAD) and MAO-mediated metabolism occupied intermediate ranks, while downstream detoxification and oxidative side reactions were comparatively minor.

In PD, the overall rank ordering of reactions was preserved, with vesicular processes remaining the highest-ranked reactions; however, their absolute rates were dramatically reduced compared to controls, reflecting the profound depletion of vesicular dopamine. As a result, dopamine synthesis (TH/LAAAD) and MAO-dependent cytosolic metabolism moved closer in magnitude to vesicular trafficking steps than in Controls, indicating a relative compression of the flux hierarchy.

According to the complete model, Control-PD differences in intra-neuronal reaction rates were, in descending order of vesicular uptake ≈ vesicular leakage > exocytotic release ≈ neuronal reuptake > L-aromatic-amino-acid decarboxylase activity ≈ tyrosine hydroxylase activity.

### Sensitivity analyses

Sensitivity analyses were performed to determine whether the rank ordering of Control–PD differences in estimated intra-neuronal reaction rates depended on assumed values of selected effective rate constants. Increasing kDA_Release over a 10-fold range (0.005–0.05 min ¹) did not affect the qualitative rank ordering (eTable 2). Decreasing kALDH by up to 90% also did not affect the rank (eTable 3).

## DISCUSSION

In this project we constructed and applied progressively more complex kinetic models for the synthesis, storage, release, reuptake, and metabolism of endogenous dopamine in putamen terminals in PD and control groups. The simplest model consisted of eight and the final model 18 reactions.

The post-mortem data demonstrated that in PD putamen DOPAL is built up with respect to dopamine—by about 9-fold. In our experience, in control subjects DOPAL is detected routinely in putamen tissue, in contrast with no detectable DOPAL in myocardium, perhaps because of rapid vesicular uptake and intra-vesicular conversion of dopamine to norepinephrine in sympathetic nerves. The availability of empirical post-mortem data about DOPAL content in putamen tissue was crucial for estimating the rates of enzymatic oxidation of DOPALc by ALDH and reduction by AR and, by applying equilibrium equations, estimating the rates of MAO_DA and VMAT_DA in putamen dopaminergic terminals.

According to the complete model, control-PD differences in intra-neuronal reaction rates were in descending order of vesicular uptake ≈ vesicular leakage > exocytotic release ≈ neuronal reuptake > L-aromatic-amino-acid decarboxylase activity ≈ tyrosine hydroxylase activity > other reactions. This rank order was qualitatively similar to that obtained from computational modeling of reactions in cardiac sympathetic nerves, where norepinephrine is the relevant neurotransmitter catecholamine,^9^ raising the possibility of shared mechanisms of catecholaminergic neurodegeneration inside and outside the brain in PD.

Sensitivity analysis for increasing kDA_Release up to 10-fold did not change the ranking, meaning that altered release with escape of reuptake was not a major contributor to putamen DA depletion. Decreasing kALDH by up to 90% also did not change the ranking, so that impaired aldehyde detoxification was not a major determinant of the increased DOPAL/DA ratio in PD putamen.

Quantitatively, estimated differences in reaction rates between controls and PD in putamen dopaminergic terminals in the present study were approximately 2-3 times slower those between controls and Lewy body diseases in sympathetic noradrenergic nerves in the heart.^9^ The higher rates in the heart suggest faster turnover of norepinephrine in cardiac sympathetic nerves than of dopamine in putamen dopaminergic terminals. In confirmation of this view, putamen ^18^F-DOPA-derived radioactivity declines from the peak value with a half-time of about 5 hours,^43^ whereas cardiac ^18^F-dopamine-derived radioactivity declines bi-exponentially from the peak value with an effective biologic late half-time on the order of 1-2 hours.^44^

Empirical in vivo data exist for the net vesicular sequestration of DAc, based on the “washout” of putamen ^18^F-DOPA-derived radioactivity.^43^ Over the course of about 70 minutes, from a static 15’ image beginning 30 minutes after initiation of tracer administration (midpoint about 38’) to a static 15’ image ending 120 minutes after initiation of tracer administration (midpoint about 108’), in controls there is about 13% decrease in radioactivity, while in PD there is about a 38.5% decrease,^43^ a difference of about 3-fold, meaning an approximately 75% decrease in net sequestration (i.e., PD retains only about 25% of control net sequestration). From the complete model, the equilibrium equations enabled separate estimations of VMAT_DA and Leak_DA, and control-PD differences were about the same for the two reactions.

In a study of the kinetics of uptake by isolated vesicles in controls and PD patients^7^ total dopamine uptake per striatal volume was reduced by 87–90% and VMAT binding by 71–80%. After correcting for dopamine nerve terminal loss, dopamine uptake per VMAT site was estimated to be reduced by 55%, while vesicular leakage was about 1.9-fold higher in PD. Thus, attenuated net sequestration seems to reflect about equal contributions of decreased vesicular uptake (about 1/2 of control) and increased passive leakage (about 2 times control), in agreement with the estimations from the complete model.

Since there were no empirical post-mortem data about DAc, DAc acted as an internal free parameter permitting back-calculation for model validation, by using the modeling data to estimate DAc and compare the estimated DAc amount with that based on available literature from cultured catecholaminergic cells.^45, 46^ In controls, estimated DAc was approximately 1.31 nmol, while empirical DAv was 446 nmoles (DAc/DAv = 0.00294). Previously reported cytoplasmic dopamine concentrations in cultured catecholamine-producing cells are approximately 1–3 µM.^9, 40^ Assuming vesicular concentrations in the 0.1–0.5 M range and vesicles comprising ∼0.3–3% of intracellular volume,^47, 48^ the expected ratio of DAc/DAv would be on the order of 0.0001–0.0100. The model-predicted DAc/DAv ratio fits squarely within the expected physiological range.

Back-calculating assuming vesicles are 1% of the volume of the terminals, vesicular dopamine is concentrated by about 34,000–fold compared to cytoplasmic dopamine, which is also in agreement with observations from cellular research.^49^ This extreme concentration gradient helps understand why even a small decrease in the efficiency of vesicular sequestration could exert large effects on the ability to maintain neurotransmitter stores.

We have previously attributed the accumulation of DOPAL with respect to dopamine in PD putamen to a combination of inefficient vesicular sequestration of cytoplasmic dopamine and decreased DOPAL detoxification by ALDH;^11^ however, the relative contributions of these determinants could not be ascertained. Based on the rankings of reaction rate abnormalities derived from the complete model, we now propose that the buildup of DOPAL with respect to dopamine in PD putamen is (a) driven predominantly by impaired vesicular sequestration, and (b) due to vesicular uptake being reduced by approximately 1/2 and vesicular leakage being approximately doubled.

### Limitations

The present results should be interpreted as steady-state, intra-neuronal estimates rather than dynamic or causal models, and the primary conclusions rely on rank ordering and relative magnitudes rather than on precise numeric values.

Since the focus of the modeling was on intra-neuronal reactions and reactants, potentially relevant extraneuronal processes such as neuroinflammation, glial cell activation, and prion-like spread of misfolded alpha-synuclein were not considered; however, none of these potentially pathogenic abnormalities can explain the drastic putamen dopamine depletion and relative DOPAL buildup that characterize PD without taking into account the intra-neuronal reactions that determine releasable dopamine stores.

Although the model is complex, it actually is overly simple, because it does not consider extraneuronal enzymatic processes such as O-methylation and sulfoconjugation,^50^ binding to catecholamine receptors, uptake into non-catecholaminergic neurons or glial cells, or effects of neuroendocrine or neuroinflammatory influences (e.g., neurosteroids, cytokines). The justification is that intra-neuronal processes were the dominant focus.

The model as constructed does not enable estimation of the relative contributions of denervation vs. functional abnormalities in residual terminals to putamen dopamine depletion. The contribution of denervation might increase as the disease progresses, but it is also possible that the two abnormalities are locked in time. More generally, because of the assumption of steady-state conditions for calculating reaction rates based on equilibrium equations, the present model does not address at all the dynamics of disease pathogenesis over years. The present model provides a platform for further extension to predict disease progression as well as effects of genetic predispositions, environmental exposures, and treatments on that progression, as has already been reported for cardiac catecholamine deficiency in Lewy body diseases.^12^

Whereas extensive literature documents putamen dopamine loss in PD, empirical published data about concentrations of most of the other analytes remain scarce. The reaction rate estimates were drawn from published point values rather than fitted to an experimental dataset, and so conventional goodness-of-fit regression statistics were not conducted.

The world’s literature on DOPAL consists of only about 165 articles, even though virtually all of the metabolism of endogenous dopamine passes through DOPAL. The catecholaldehyde hypothesis has never been tested formally in humans. Nevertheless, to date all cellular and animal models of PD that have been examined for striatal DOPAL content—and clinical PD—have been associated with endogenous DOPAL buildup with respect to dopamine, and all manipulations known to increase generation of endogenous DOPAL have been associated with motor abnormalities resembling those in PD.^3, 14, 51–60^

### Implications

Neither nigrostriatal dopaminergic denervation nor reduced dopamine synthesis in putamen dopamine terminals can account for the empirical observation of an increased DOPAL/DA ratio in PD. The estimated reaction rates generated from the complete model highlight functional abnormalities that lead to impaired vesicular sequestration of dopamine in extant terminals. The findings challenge the sufficiency of nigrostriatal dopaminergic denervation alone to account for the biochemical phenotype of PD and highlight vesicular dopamine handling as a critical determinant of putamen dopamine deficiency. The present modeling approach identifies specific targets such as VMAT2 and vesicular leakage for disease-modifying treatments that might delay the onset of symptomatic PD by mitigating the loss of putamen dopamine.

## Supporting information

eData 1-Autopsy Individual Data No Ages

eTable 1, Reactants in Literature.

eTable 2 Sensitivity_kDA_Release

eTable 3 Sensitivity_kALDH

## Data Availability

All data produced in the present study are available upon reasonable request to the author

## Author contributions

Dr. Goldstein devised the project, culled and analyzed the data, generated the Figures, and wrote the manuscript.

The author retired from federal employment in August, 2025 and now is a Scientist Emeritus. The contributions of the author were made as part of official duties when he was an NIH federal employee, are in compliance with agency policy requirements, and are considered Works of the United States Government.

The findings and conclusions presented in this paper are those of the author and do not necessarily reflect the views of the NIH or the U.S. Department of Health and Human Services.

## Acknowledgements

Courtney Holmes, CMT, and Patti Sullivan, CMT, assayed the tissue samples referred to in this project. Drs. Irwin J. Kopin (deceased), Yehonatan Sharabi, Mark Pekker, and Graeme Eisenhofer collaborated on previous studies involving computational modeling that inspired the present project.

## Acknowledgement statement (including conflict of interest and funding sources)

This research was supported [in part] by the Intramural Research Program (ZIA NS003034) of the National Institutes of Health (NIH), National Institute of Neurological Disorders and Stroke (NINDS).

In the past 12 months David S. Goldstein established The Autonomic and Catecholamine Healthspan Institute, LLC. He continues to receive an annual royalty for a book published by Johns Hopkins University Press in 2006. The author declares that there are no additional disclosures to report.

---

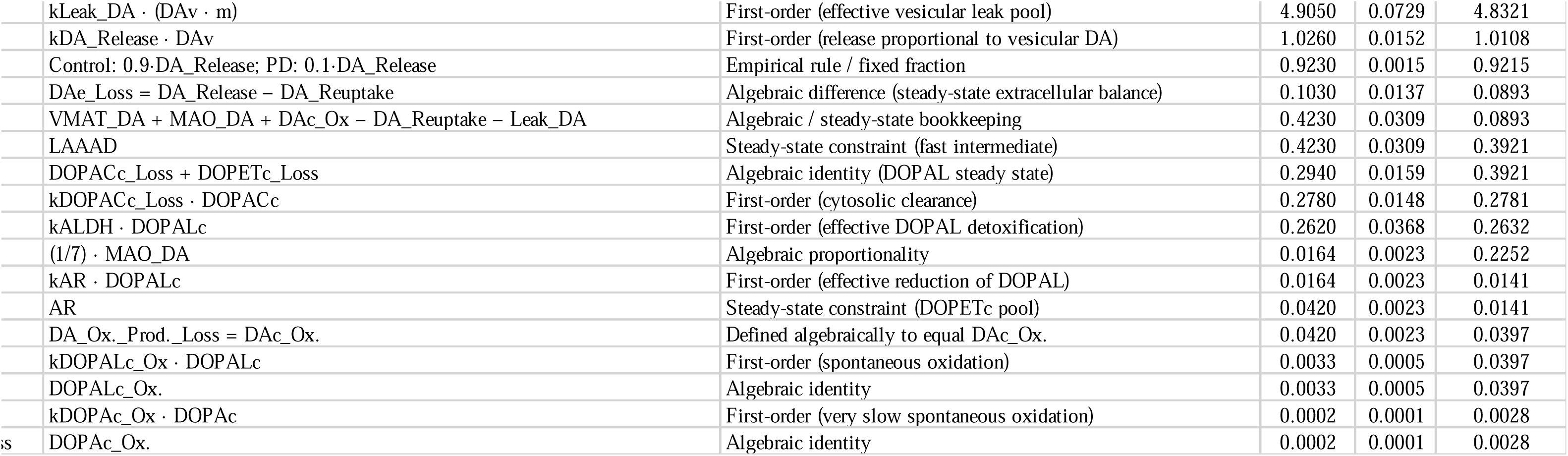

